# Functional Shotgun Metagenomic Insights into Gut Microbial Pathway and Enzyme Disruptions Linking Metabolism, Affect, Cognition, and Suicidal Ideation in Major Depressive Disorder

**DOI:** 10.64898/2025.12.10.25342027

**Authors:** Michael Maes, Abbas F. Almulla, Asara Vasupanrajit, Ketsupar Jirakran, Chavit Tunvirachaisakul, Annabel Maes, Prangwalai Chancham, Pavit Klomkliew, Sunchai Payungporn, Yingqian Zhang

## Abstract

**Background:** Major depression (MDD) is linked to neuro-immune, metabolic, and oxidative stress (NIMETOX) pathways. The gut microbiome may contribute to these pathways via leaky gut and immune-metabolic processes.

**Aims:** To identify gut microbial alterations in MDD and to quantify functional pathways and enzyme gene families and integrate these with the clinical phenome and immune–metabolic biomarkers of MDD.

**Methods:** Shotgun metagenomics with **t**axonomic profiling was performed in MDD versus controls using MetaPhlAn v4.0.6, and functional profiling was conducted using HUMAnN v3.9, aligning microbial reads to species-specific pangenomes (Bowtie2 v2.5.4) followed by alignment to the UniRef90 v201901 protein database (DIAMOND v2.1.9).

**Results:** Gut microbiome diversity, both species richness and evenness, is quite similar between MDD and controls. The top enriched taxa in the multivariate discriminant profile of MDD reflect gut dysbiosis associated with leaky gut and NIMETOX mechanisms, i.e., *Ruminococcus gnavus, Veillonella rogosaem* and *Anaerobutyricum hallii*. The top four protective taxa enriched in controls indicate an anti-inflammatory ecosystem and microbiome resilience, i.e., *Vescimonas coprocola*, C*oprococcus*, *Faecalibacterium prausnitzii*, and *Faecalibacterium parasitized*. Pathway analysis indicates loss of barrier protection, antioxidants and short-chain fatty acids, and activation of NIMETOX pathways. The differential abundance of gene families suggests that there are metabolic distinctions between both groups, indicating aberrations in purine, sugar, and protein metabolism. The gene and pathway scores explain a larger part of the variance in suicidal ideation, recurrence of illness, neurocognitive impairments, immune functions, and atherogenicity.

**Conclusion:** The gut microbiome changes might contribute to activated peripheral NIMETOX pathways in MDD.

## Introduction

Evidence currently indicates that major depressive disorder (MDD) is linked to neuro-immune, metabolic, and oxidative stress (NIMETOX) pathways (Maes, Almulla, et al., 2025; Maes, Jirakran, et al., 2025). Various factors, such as viral infections, adverse childhood experiences (ACEs), negative life events, and heightened translocation of Gram-negative bacteria, may activate the NIMETOX pathways, potentially resulting in enhanced neurotoxicity to affective circuits in the brain (Maes, Almulla, et al., 2025; Maes, Jirakran, et al., 2025). The neuro-immune pathways encompass neurotoxic inflammatory cytokines, such as those of the M1 macrophage and T helper (Th)1 lineage, and a disbalance in growth factors, including vascular endothelial growth factor (VEGF) and stem cell factor (SCF, kit ligand) (Maes et al., 2024). The VEGF/SCF ratio is elevated in MDD, indicating diminished endothelial resilience and increased microvascular permeability (Maes, Zhou et al. 2024). VEGF is stimulated by cytokine-dependent pathways (e.g., IL-1β, TNF-α) and facilitates leukocyte trafficking and angiogenesis, and increases vascular permeability (Angelo & Kurzrock, 2007). SCF/c-KIT signaling maintains hematopoietic stem/progenitor reservoirs, mast cell-mediated vascular control, and structured tissue repair (Broudy, 1997).

The principal metabolic aberrations in MDD include heightened atherogenicity characterized by diminished levels of high-density lipoprotein cholesterol (HDL) and elevated proatherogenic lipid fractions such as free cholesterol, triglycerides, and apolipoprotein B (ApoB) (Chen et al., 2025; Maes et al., 1994). Elevated lipid peroxidation and oxidative damage to proteins, coupled with diminished antioxidant defenses, represent additional mechanisms that, with neuro-immune and metabolic pathways, may contribute to heightened neurotoxicity (Maes, Almulla, et al., 2025; Maes, Jirakran, et al., 2025). Moreover, these NIMETOX pathways significantly influence the clinical phenome (representing a comprehensive index of observable clinical characteristics of MDD), encompassing the overall severity of depression (OSOD), recurrence of illness (ROI), and suicidal behaviors (Maes, Almulla, et al., 2025; Maes, Jirakran, et al., 2025; Maes et al., 2024).

Evidence suggests that disturbances in the gut microbiome may activate NIMETOX pathways through enhanced translocation of endotoxin (LPS) from the gut to the peripheral blood, a phenomenon referred to as leaky gut or increased gut permeability (Maes et al., 2008; Rudzki & Maes, 2020; Rudzki et al., 2021). Moreover, gut dysbiosis in MDD frequently manifests as an increase in pathobionts, which exacerbate inflammatory responses and intestinal permeability, alongside a reduction in taxa that synthesize protective short-chain fatty acids (SCFAs), which enhance tight junctions and mucosal immunity, consequently facilitating leaky gut and LPS translocation (Cheung et al., 2019; Rudzki et al., 2021). Furthermore, gut dysbiosis modifies the synthesis of neurotransmitters and the production and availability of microbial vitamins (e.g., folate, vitamins B , B , B , B ) that are essential for one-carbon metabolism associated with mood regulation (Magnúsdóttir et al., 2015; Radjabzadeh et al., 2022; Rudzki et al., 2021).

Amplicon sequencing studies of 16S ribosomal RNA (rRNA) in MDD compared to controls reveal mild and varied compositional alterations (Cheung et al., 2019; Gao et al., 2023; Maes, Vasupanrajit, Jirakran, Klomkliew, Chanchaem, Tunvirachaisakul, & Payungporn, 2023; Sanada et al., 2020). Variations in alpha-diversity are inconsistent; aggregated effects are minimal or negligible, with area and medication contributing to variability (Maes, Vasupanrajit, Jirakran, Klomkliew, Chanchaem, Tunvirachaisakul, & Payungporn, 2023; Nikolova et al., 2021; Sanada et al., 2020). Beta diversity frequently varies among groups, though not consistently (Cheung et al., 2019; Maes, Vasupanrajit, Jirakran, Klomkliew, Chanchaem, Tunvirachaisakul, & Payungporn, 2023; Sanada et al., 2020). Frequently diminished genera in MDD that produce beneficial SCFAs include *Ruminococcus, Faecalibacterium,* and *Coprococcus* (Gao et al., 2023; Maes, Vasupanrajit, Jirakran, Klomkliew, Chanchaem, Tunvirachaisakul, & Payungporn, 2023; Nikolova et al., 2021). Conversely, MDD is characterized by pro-inflammatory taxa, mucin-utilizers and pathobionts such as *Hungatella, Clostridium bolteae,* and *Hungatella hathewayi, Eggerthella* (Maes, Vasupanrajit, Jirakran, Klomkliew, Chanchaem, Tunvirachaisakul, & Payungporn, 2023; Radjabzadeh et al., 2022; Sanada et al., 2020). Employing the 16S rRNA amplicon sequencing technique, Maes et al. (Maes, Vasupanrajit, Jirakran, Klomkliew, Chanchaem, Tunvirachaisakul, Plaimas, et al., 2023) identified two “enterotypes” (composites of enriched pathobionts and depleted symbionts), with the first elucidating a significant portion of the variance in the phenome of MDD including suicidal tendencies, and the second accounting for a greater extent of the variance in metabolic variables, such as HDL cholesterol.

Amplicon sequencing focuses on 16S rRNA, utilizing PCR primers that attach to regions adjacent to hypervariable segments. This approach provides resolution at the genus and species level; however, it does not facilitate direct quantification of genes or pathways (Klindworth et al., 2013). Moreover, the performance is contingent upon the selection of primers, which can lead to amplification bias, even when utilizing validated primer sets (Klindworth et al., 2013; Tremblay et al., 2015). The estimates of abundance are additionally influenced by the variability in 16S copy numbers among different taxa (Johnson et al., 2019). Conversely, shotgun metagenomics offers a comprehensive, PCR-free approach to genome-wide profiling across various domains, such as bacteria and archaea, achieving species and strain resolution while enabling direct quantification of genes and pathways (Quince et al., 2017). This approach effectively circumvents primer-mismatch bias and the distortion of marker-gene copy numbers, both of which can significantly impact amplicon studies (Johnson et al., 2019; Klindworth et al., 2013). Contemporary workflows, exemplified by HUMAnN, facilitate the mapping of reads to gene families while reconstructing pathway abundance and coverage. This process allows for the establishment of mechanistic connections between microbiome taxa and their corresponding functions (Franzosa et al., 2018).

A shotgun study contrasting MDD and bipolar depression revealed diagnosis-associated variations in microbial abundances (Rong et al., 2019). Another case–control shotgun study in MDD has reported alterations in community structure, characterized by shifts in tryptophan biosynthesis and degradation, thereby establishing a connection between taxonomy and serotonergic metabolism (Lai et al., 2021). In conclusion, advancing from mere association to a deeper understanding of mechanisms, as well as enhancing reproducibility in the realm of MDD-microbiome research, necessitates the implementation of additional high-quality shotgun metagenomic studies.

Hence, the objective of this pilot shotgun metagenomics study is to (1) identify microbial alterations at the species and strain levels in MDD; (2) quantify differences in functional pathways and gene enzymes; (3) integrate microbial functions (pathways and genes) with immune–metabolic biomarkers and the clinical phenome of MDD; and (4) delineate the pathway and gene signatures that predict the MDD phenome.

## Methods

### Participants

We recruited 37 normal controls and 32 MDD patients from the outpatient clinic of the Department of Psychiatry at King Chulalongkorn Memorial Hospital in Bangkok, Thailand (Maes et al., 2024). In part of these patients (n = 23) and controls (n = 23), we performed shotgun metagenomics analysis. Participants ranged in age from 19 to 58 and were of both sexes. MDD was diagnosed in patients who were in an acute depressive episode using the DSM-5 criteria.

### Clinical assessments

A research psychologist experienced in affective disorders conducted semi-structured interviews to collect socio-demographic and clinical data. A senior psychiatrist diagnosed MDD per DSM-5 criteria using the Thai-version M.I.N.I. (Udomratn & Kittirattanapaiboon et al., 2004) for Axis I assessment and participant exclusion. Assessments included HAMD, Thai-translated BDI-II (Thavichachart et al., 2009), STAI, C-SSRS (Posner et al., 2011; deriving OSOD/ROI per Maes et al., 2024), Stroop test (z composite), Thai ACEs questionnaire (5 domains; z score + sexual abuse score, Rungmueanporn et al., 2019). MetS (IDF criteria), BMI (weight/height²), and TUD (DSM-5) were evaluated.

### Assays

#### DNA Extraction and shotgun metagenomic sequencing

DNA was extracted from fecal samples (approximately 20 mg) using ZymoBIOMICS™ DNA Miniprep Kit (ZYMO Research, USA) following the manufacturer’s protocol. Then, 100 ng of extracted DNA were used for DNA library construction using MGIEasy FS DNA Library Prep Set (MGI, China) according to the manufacturer’s recommended protocol. After that, DNA libraries were sequenced with paired-end (2 x 150 bp) using the DNBSEQ-G400 platform (MGI, China) following the manufacturer’s recommendations.

#### Metagenomic Sequence Data Processing and Differential Abundance Analyses

Raw paired-end reads underwent FastQC v0.11.9 quality control (GC content, base quality, adapter contamination), followed by Fastp v0.20.1 trimming (Phred score <30), adapter removal, and discarding reads <60 bp. Human DNA was eliminated via BWA v0.7.18 alignment to hg38; Samtools v1.20 filtered out human reads, retaining microbial data.

Taxonomic profiling used MetaPhlAn v4.0.6 (mpa_vJun23_CHOCOPhlAnSGB_202307 database, clade-specific marker genes), keeping species with >0.1% relative abundance in ≥5 samples. Functional profiling employed HUMAnN v3.9: microbial reads aligned to species pangenomes via Bowtie2 v2.5.4 and to UniRef90 v201901 via DIAMOND v2.1.9.

#### Statistics

Functional abundances (gene families/pathways) were normalized to RPK and CLR-transformed. Microbiota/taxonomic analyses used vegan R package; alpha diversity (Shannon/richness) was compared via Mann-Whitney U test, beta diversity via Bray-Curtis PCoA. Hierarchical clustering (hclust2) and Random Forest (caret/randomForest) classified MDD/HC groups, with Ridge regression (SIAMCAT) identifying discriminatory gene families. Pathway differences were visualized via ggplot2, and Spearman networks via igraph/ggraph. Categorical variable associations used Chi-square tests, continuous variables ANOVA. Automated/manual regression (Statistica/SPSS) identified predictors for phenome/biomarkers, with model checks for normality, collinearity, and homoscedasticity. Sample size (n=30) was calculated via G*Power 3.1.9.4, based on previously study(Maes et al., 2024), with 0.8 power, α=0.05, and effect size explaining ∼35% phenome variance. All statistical analyses (IBM SPSS 30) used two-tailed *p* < 0 05. See more details in **Supplementary methods.**

## Results

### Demographic and clinical data

**Table 1** lists the clinical and demographic data of the participants. There were no significant differences in age, sex ratio, smoking, metabolic variables, education between both study samples. Clinical scores (OSOD, SI, Stroop score, ROI, and ACEs) were significantly different between both groups. The composite score of VEGF – SCF was significantly higher in patients than controls but there were no significant differences in AIP.

**Table 1.** Demographic and clinical data in patients with major depression (MDD) and healthy controls (HC).

| Variables | HC (n = 23) | MDD (n = 23) | F / chi-square | df | *p* |
| --- | --- | --- | --- | --- | --- |
| Age (years) | 29.5 (7.7) | 25.7 (8.0) | 2.67 | 1/44 | 0.109 |
| Female/male | 16/4 | 18/5 | 0.14 | 1 | 0.710 |
| Normal BMI/overweight/obesity | 19/2/2 | 17/4/2 | 0.78 | 2 | 0.678 |
| Metabolic syndrome (no/yes) | 20/3 | 20/3 | 0.00 | 1 | 1.0 |
| Smoking (no/yes) | 21/2 | 17/6 | 2.42 | 1 | 0.120 |
| Education (years) | 16.3 (2.2) | 16.5 (3.5) | 0.06 | 1/44 | 0.803 |
| OSOD (z score) | -0.702 (0.188) | 0.873 (0.965) | 58.91 | 1/44 | < 0.001 |
| Current SI (z score) | -0.700 (0.0) | 0.651 (0.948) | MWUT | - | < 0.001 |
| Recurrence of illness index (rank) | 0.290 (0.115) | 0.761 (0.141) | 151.22 | 1/44 | < 0.001 |
| Stroop Composite (z score) | 0.227 (0.673) | -0.357 (1.064) | 5.84 | 1/44 | 0.020 |
| ACE: neglect and abuse (z score) | -0.418 (0.903) | 0.490 (0.752) | 13.75 | 1/44 | < 0.001 |
| ACE: Sexual abuse (z score) | -0.343 (0.0) | 0.512 (1.526) | MWUT | - | < 0.001 |
| Atherogenic index of plasma (z score) | -0.116 (1.050) | 0.017 (0.906) | 0.21 | 1/44 | 0.648 |
| Growth factor profile (z score) | -0.453 (0.973) | 0.453 (0.818) | 11.66 | 1/44 | 0.001 |

All data are shown as mean ( ± SD). All results of analysis of variance, analysis of contingency tables or MWUT: Mann-Whiney U test. BMI: body mass index, OSOD: overall severity of illness, SI: suicidal ideation, ACE: adverse childhood experiences.

### Microbial Diversity and Composition

**Figure 1 (A-C)** presents a comparative analysis of microbial diversity and beta diversity between the control (HC) and MDD groups. In **Figure 1A**, the observed species richness, reflecting the total number of distinct microbial species present, indicates that the control group has a slightly higher average species richness compared to the MDD group. However, this difference is not statistically significant (*p* = 0.36), suggesting that overall microbial richness is relatively similar between the two groups. **Figure 1B** shows the Shannon diversity index, which accounts for both species richness and evenness. This index did not show a significant difference between both groups (*p* = 0.27).

**Figure 1.**
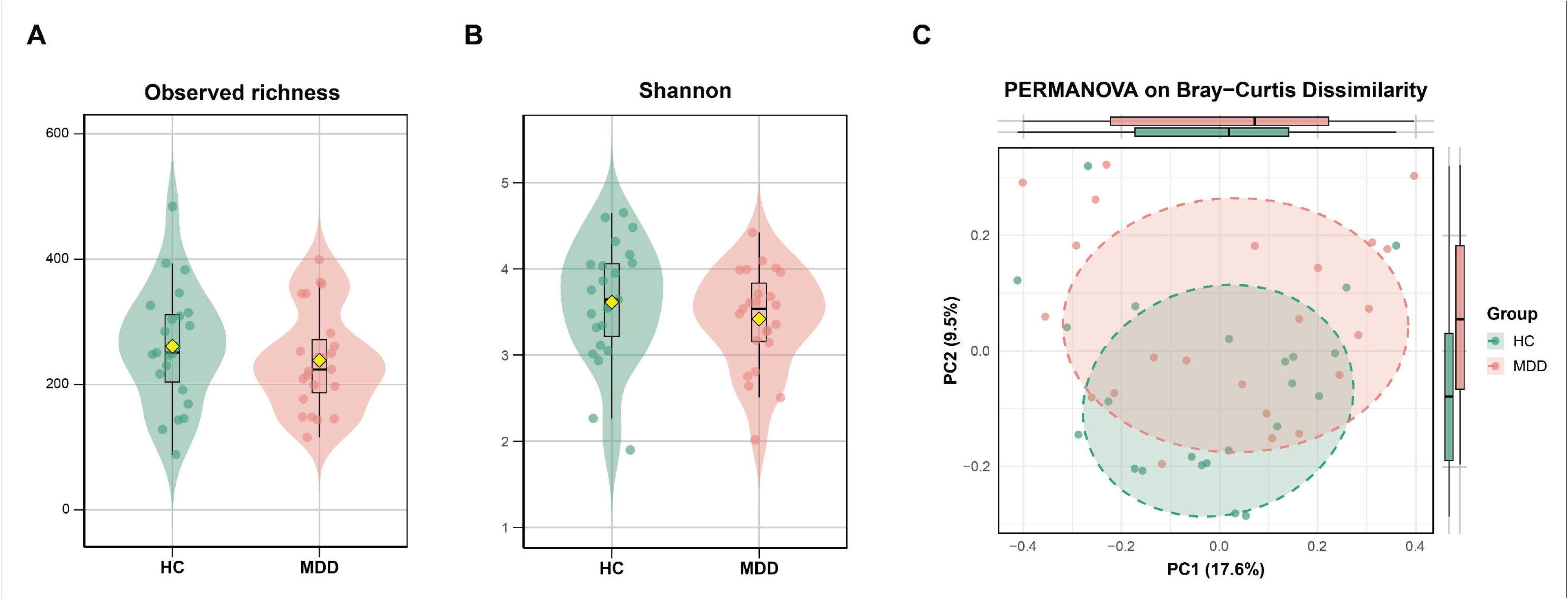
This figure presents three aspects of microbial diversity in HC and MDD groups. (A) The observed richness plot depicts the total number of distinct bacterial species detected per sample. (B) The Shannon diversity index highlights both species richness and evenness. (C) The PCoA plot visualizes microbial community composition, where the proximity of points reflects the similarity between individual samples.

In **Figure 1C**, Principal Coordinates Analysis (PCoA) based on Bray-Curtis dissimilarity is used to assess differences in microbial community composition (beta diversity) between the two groups. The PCoA plot reveals some compositional differences between their gut microbiomes. However, the overlap between the groups and the p-value from the PERMANOVA test (*p* = 0.2) shows that these differences are not statistically significant.

### Taxonomic Composition and Clustering

**Figure 2** presents a heatmap displaying the relative abundance of the top 40 most abundant bacterial species across the MDD and control groups. Hierarchical clustering was performed on both the bacterial species (rows) and individual samples (columns) to detect potential patterns in microbial composition between the groups. The heatmap shows no clear separation between both groups, with species abundances distributed across both groups, indicating that the overall taxonomic composition of the gut microbiome is quite similar between the two groups. This lack of distinct clustering suggests that there are no dominant bacterial taxa that exclusively characterize either group. Several bacterial species, such as *Faecalibacterium prausnitzii*, *Roseburia faecis*, and species from the *Bacteroides* genus, are prevalent in both groups, which reinforces the observation that there are no major taxonomic differences.

**Figure 2.**
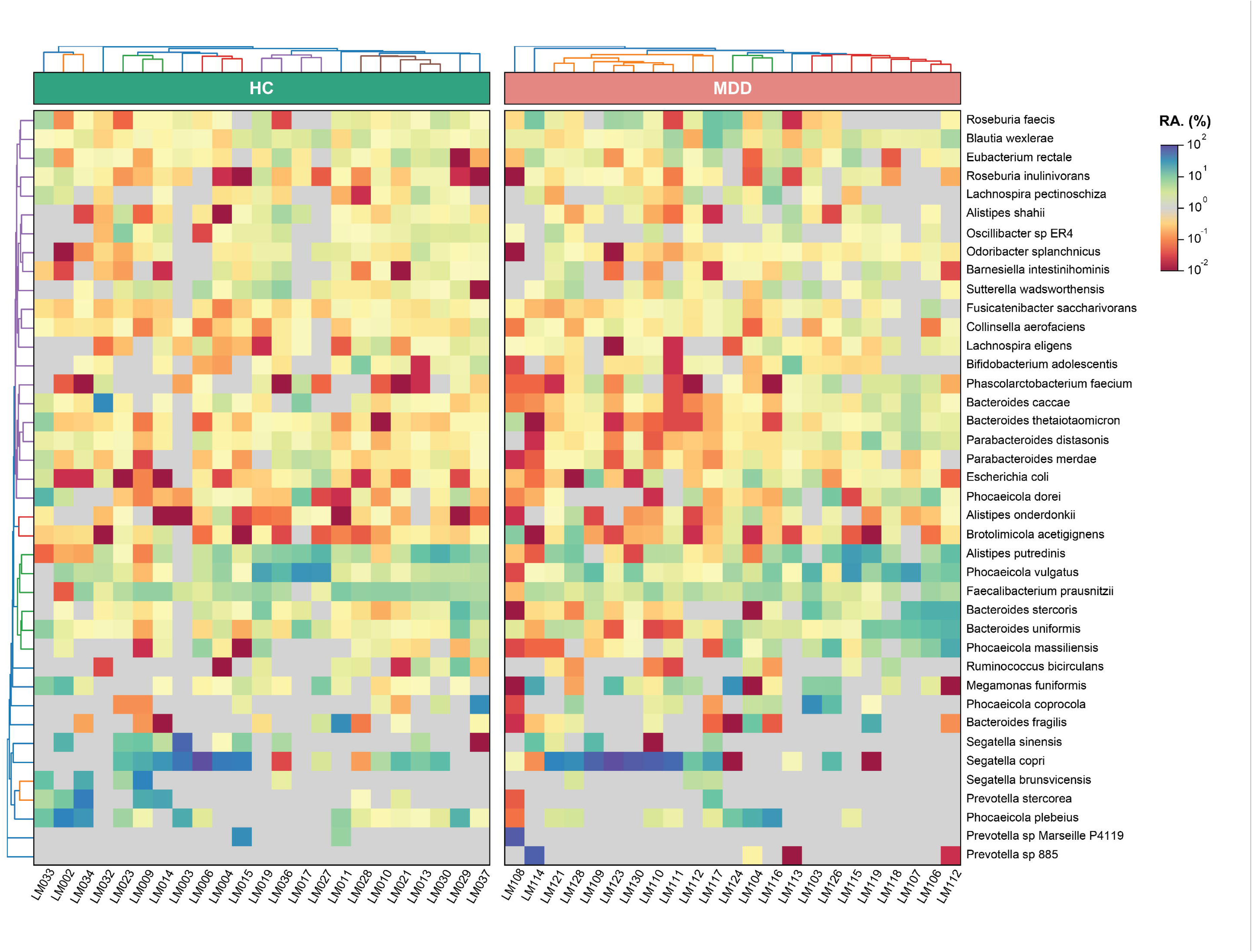
The heatmap illustrates the relative abundance of the top 40 most abundant bacterial species across both HC and MDD groups. Rows correspond to bacterial species, while columns represent individual samples. Color intensity indicates species abundance, and the cladogram on the left indicates clustering based on similarity patterns.

### Random Forest Classification of Microbial Features

To identify specific bacterial taxa that may act as biomarkers for distinguishing HC from MDD, a Random Forest classification model was applied. **Figure 3** shows the top 15 bacterial species ranked by their importance in differentiating MDD and controls. Among the species enriched in the MDD group, *Ruminococcus gnavus*, *Veillonella rogosae*, and *Anaerobutyricum hallii* were identified as significant contributors to the classification model. Conversely, species such as *Vescimonas coprocola* and *Faecalibacterium prausnitzii* were more prevalent in the control group. Nevertheless, the ROC curve achieved an Area Under the Curve (AUC) of 0.62 only, which indicates limited discriminatory power.

**Figure 3.**
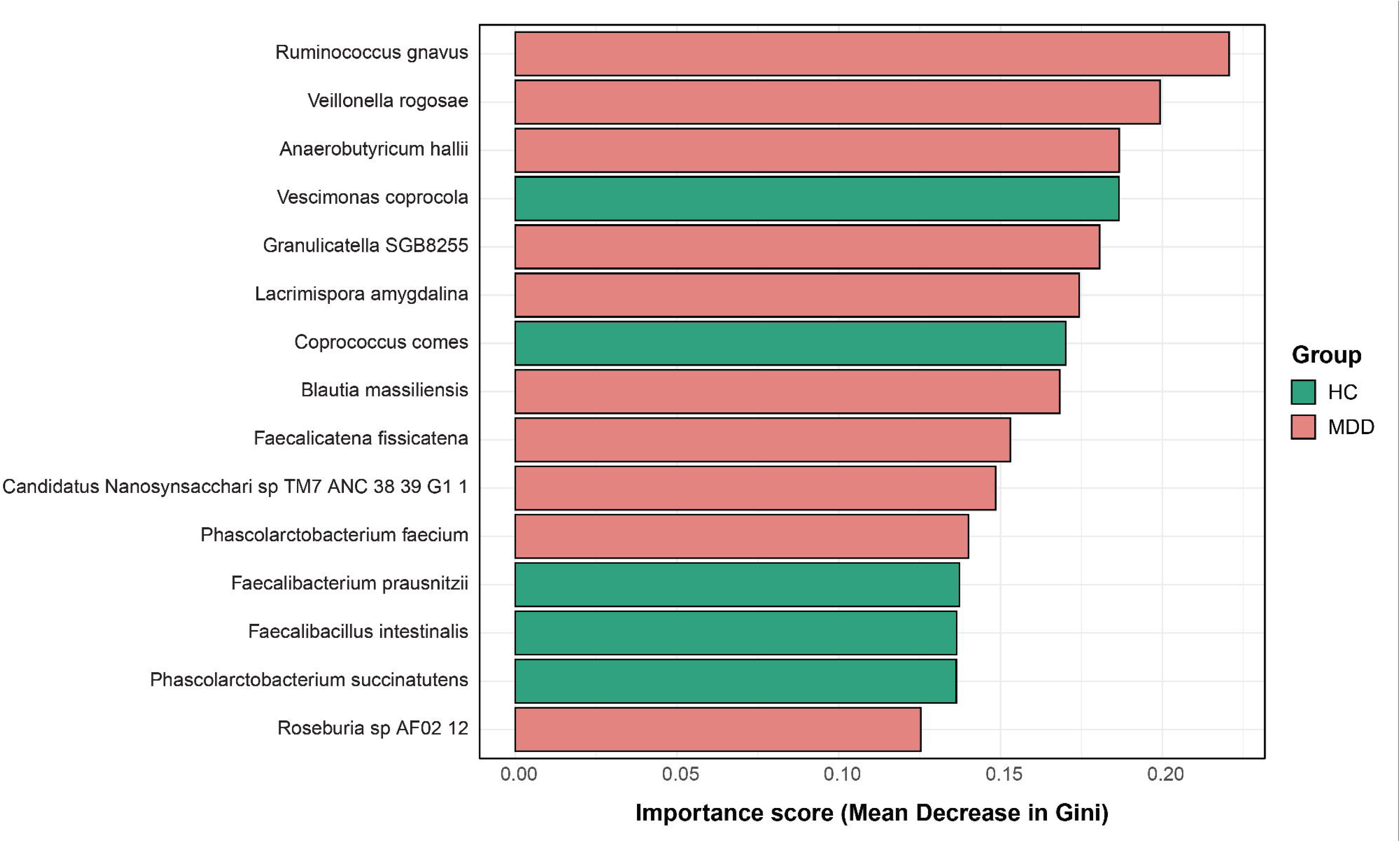
This bar plot displays the 15 most important bacterial taxa identified by the Random Forest classifier in differentiating between HC and MDD groups. Higher bars indicate taxa that contributed more to the classification, identifying microbial markers potentially associated with the MDD condition.

### Pathway Analysis and Correlation Network

The log2 fold change analysis (performed using pseudocounts) (**Figure 4**) identifies the top 20 pathways distinguishing the microbiomes of both groups. The MDD group shows enrichment in pathways such as folate transformations I, peptidoglycan biosynthesis IV, and formaldehyde assimilation. In the control group, notably enriched pathways include the superpathway of mycolate biosynthesis and 6-hydroxymethyl-dihydropterin diphosphate biosynthesis II.

**Figure 4.**
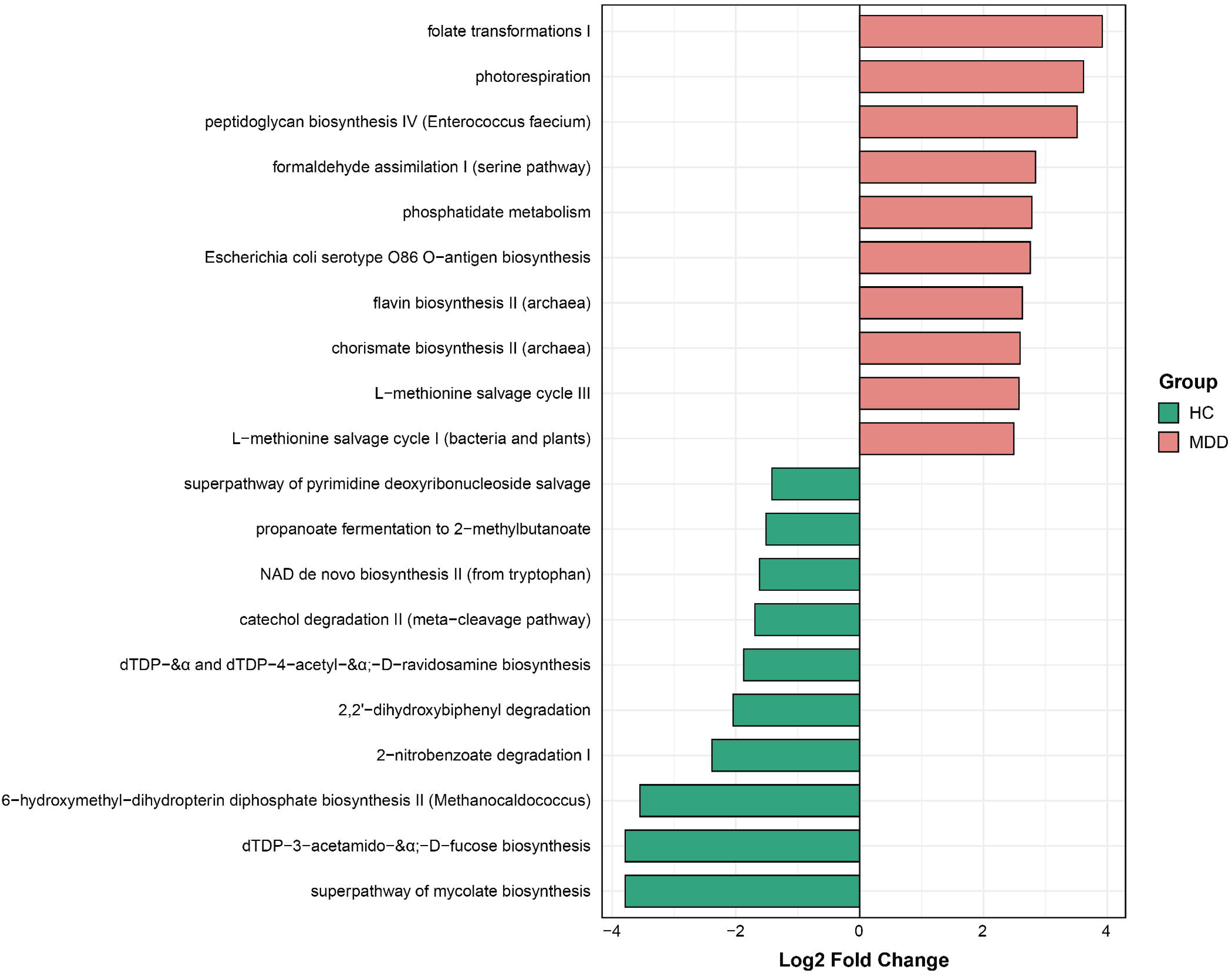
This bar plot highlights the top 20 pathways with significant differences in abundance between HC and MDD groups. The length of the bars reflects the fold change in pathway abundance, with green indicating pathways enriched in HC and red indicating pathways enriched in MDD.

The correlation network analysis (**Figure 5**) provides a deeper look at the interconnections between these pathways. The control network reveals pathways like catechol degradation II, L-tyrosine and UDP-N-acetyl-D-glucosamine biosynthesis, and gluconeogenesis as central hubs. In contrast, the MDD network indicates pathways such as LPS biosynthesis, archaetidylinositol & CDP-archaeol biosynthesis, and 7-(3-amino-3-carboxypropyl)-tyrosine biosynthesis emerging as central nodes.

**Figure 5.**
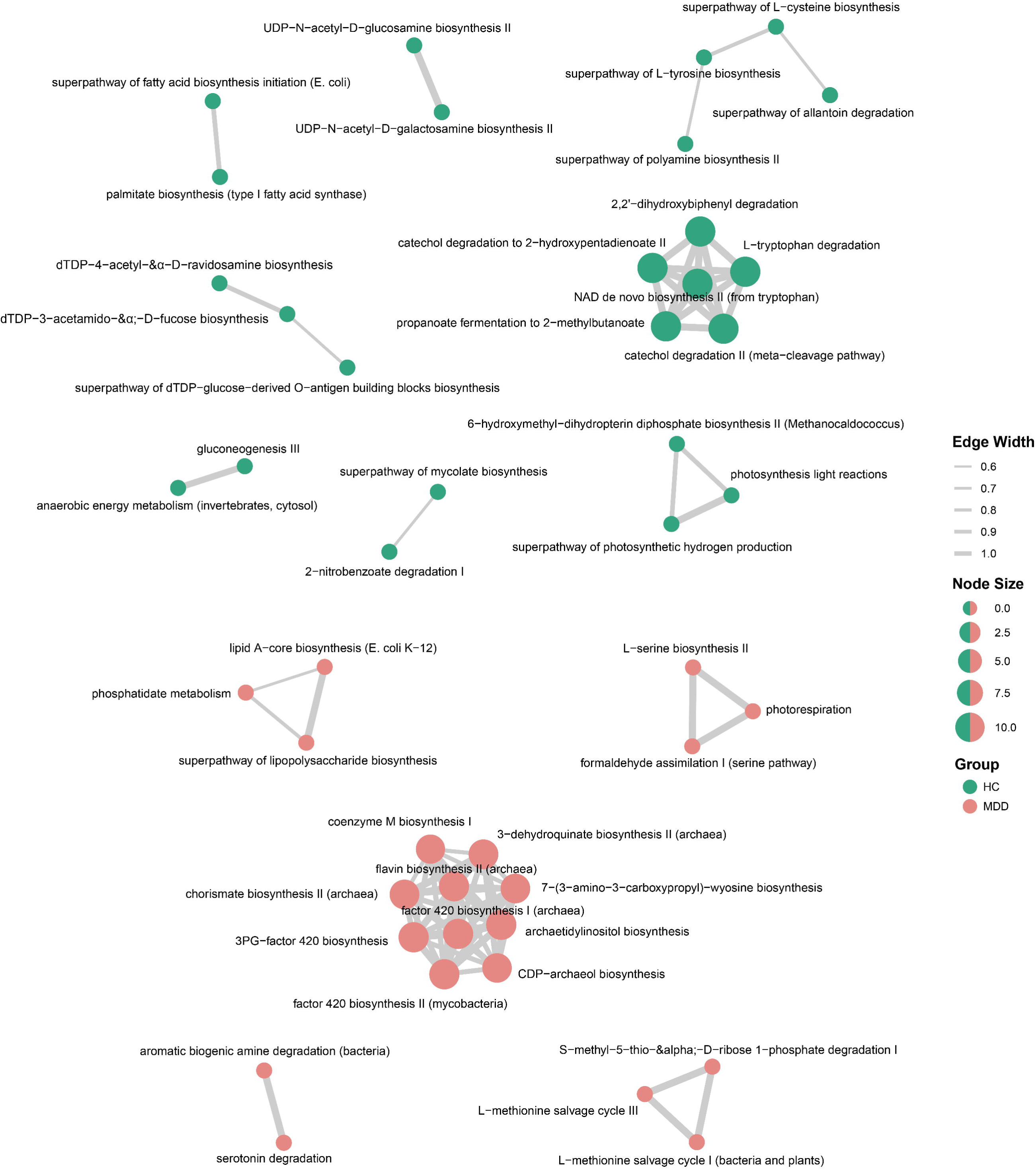
The correlation network illustrates the relationships between the top 30 pathways in both HC and MDD groups. Each node represents a pathway, with larger node sizes indicating more central pathways in the network. Pathways are connected by edges that signify significant correlations, with the thickness of the edges reflecting the strength of these relationships. The HC network (top) and the MDD network (bottom) highlight distinct pathway interactions between the two groups.

### Functional Gene Family Profiling and Classification

Functional gene family profiling was performed using a Ridge regression model to further investigate differences between MDD and controls at the functional level. **Figure 6** presents the top 20 gene families that contributed to the classification of the two groups. The gene families that were more abundant in the MDD group include Ketol-acid reductoisomerase [1.1.1.86], Dephospho-CoA kinase [2.7.1.24], and Aspartate 1-decarboxylase [4.1.1.11]. On the other hand, gene families like Glucosamine-6-phosphate deaminase [3.5.99.6] and Deoxyribose-phosphate aldolase [4.1.2.4] were found to be more abundant in the HC group. Nevertheless, the ROC curve showed a moderate AUC of 0.67.

**Figure 6.**
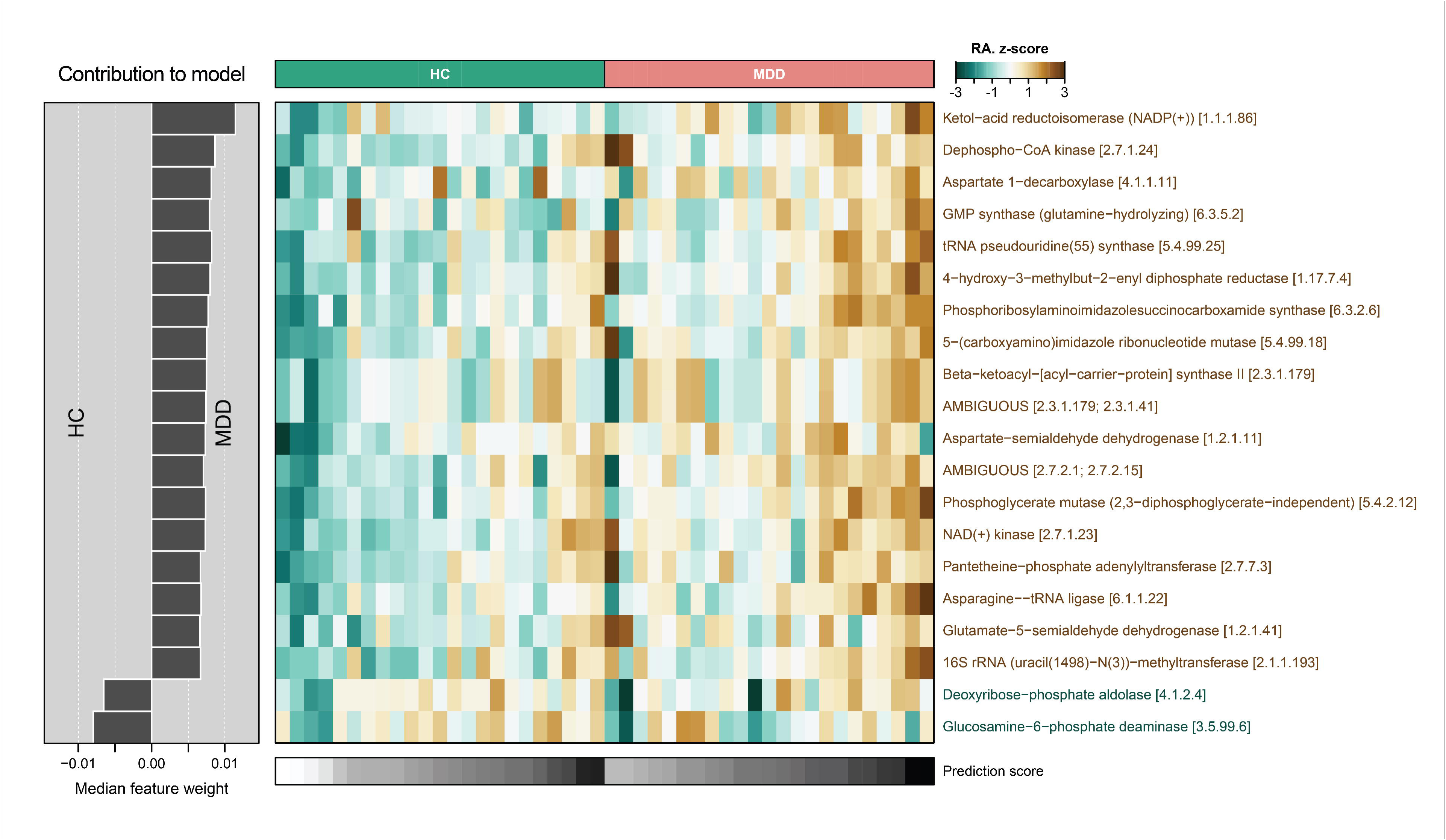
This figure presents the top 20 gene families contributing to the classification between HC and MDD. The bar plot on the left shows the feature weights, with positive values linked to MDD and negative ones to HC. The heatmap on the right visualizes the relative abundance of these gene families, with color intensity reflecting their abundance across samples. Each gene family is associated with a KEGG enzyme ID, and the legend indicates the range of prediction scores.

### Results of multiple regression analysis

**Tables 2 and 3** show the results of multiple regression analyses with clinical or metabolic biomarkers as dependent variables and either the pathways (**Table 2**) or genes **(Table 3)** as explanatory variables. Table 2, regression #1 shows that 29.8% of the variance in OSOD was explained by PWY 5989 (positively) and PWY 7254 (inversely). We found that (**Table 3**, regression #1) 54.2% of the variance in OSOD was explained by EC 6.3.5.2 and EC 5.4.99.18 (both positively) and EC 1.5.1. and EC 3.5.99.6 (both negatively).

**Table 2.**
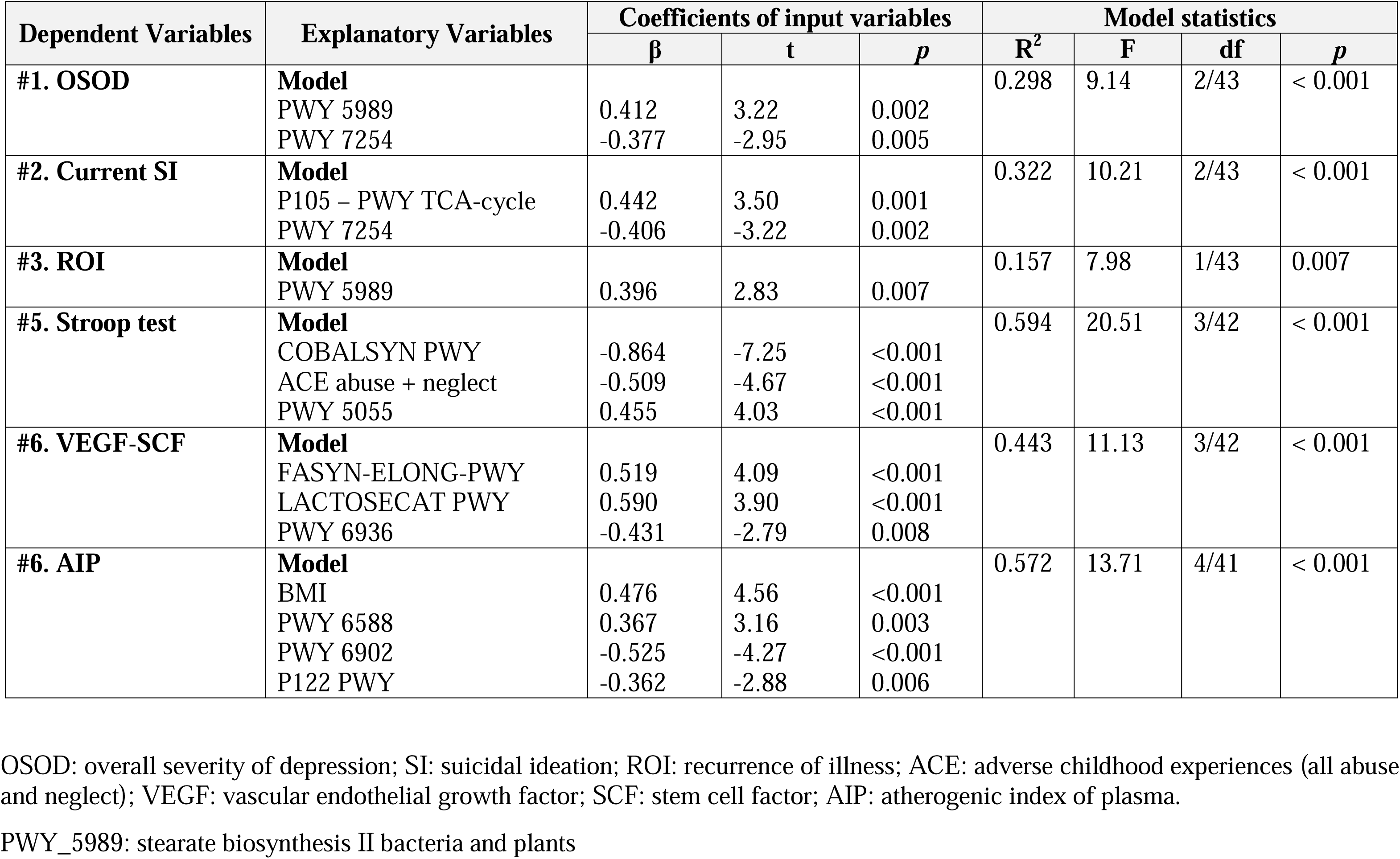
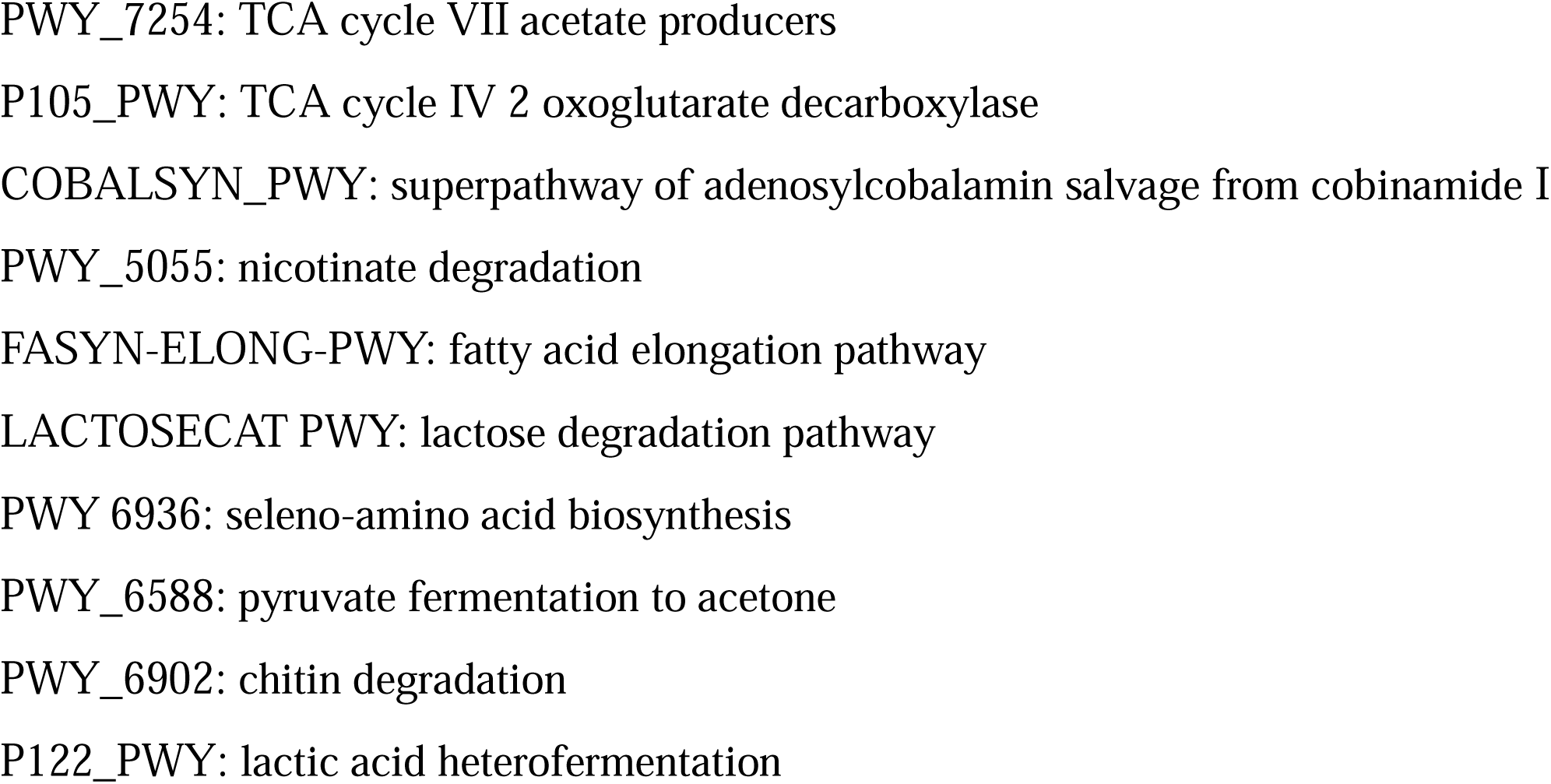
Results of multiple regression analysis with clinical data, cognitive tests scores, and biomarkers as dependent variables, and pathway scores as explanatory variables.

**Table 3.**
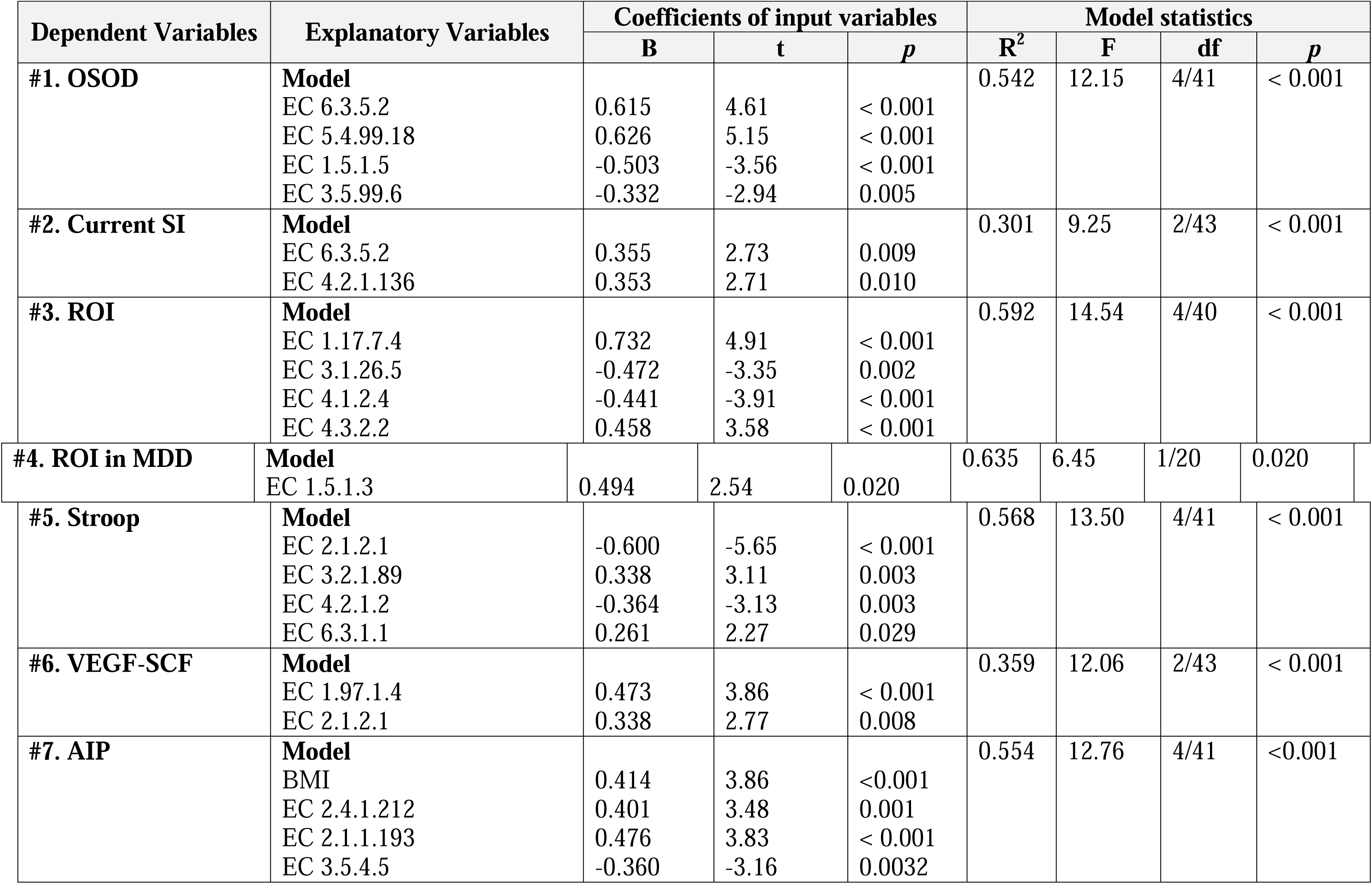
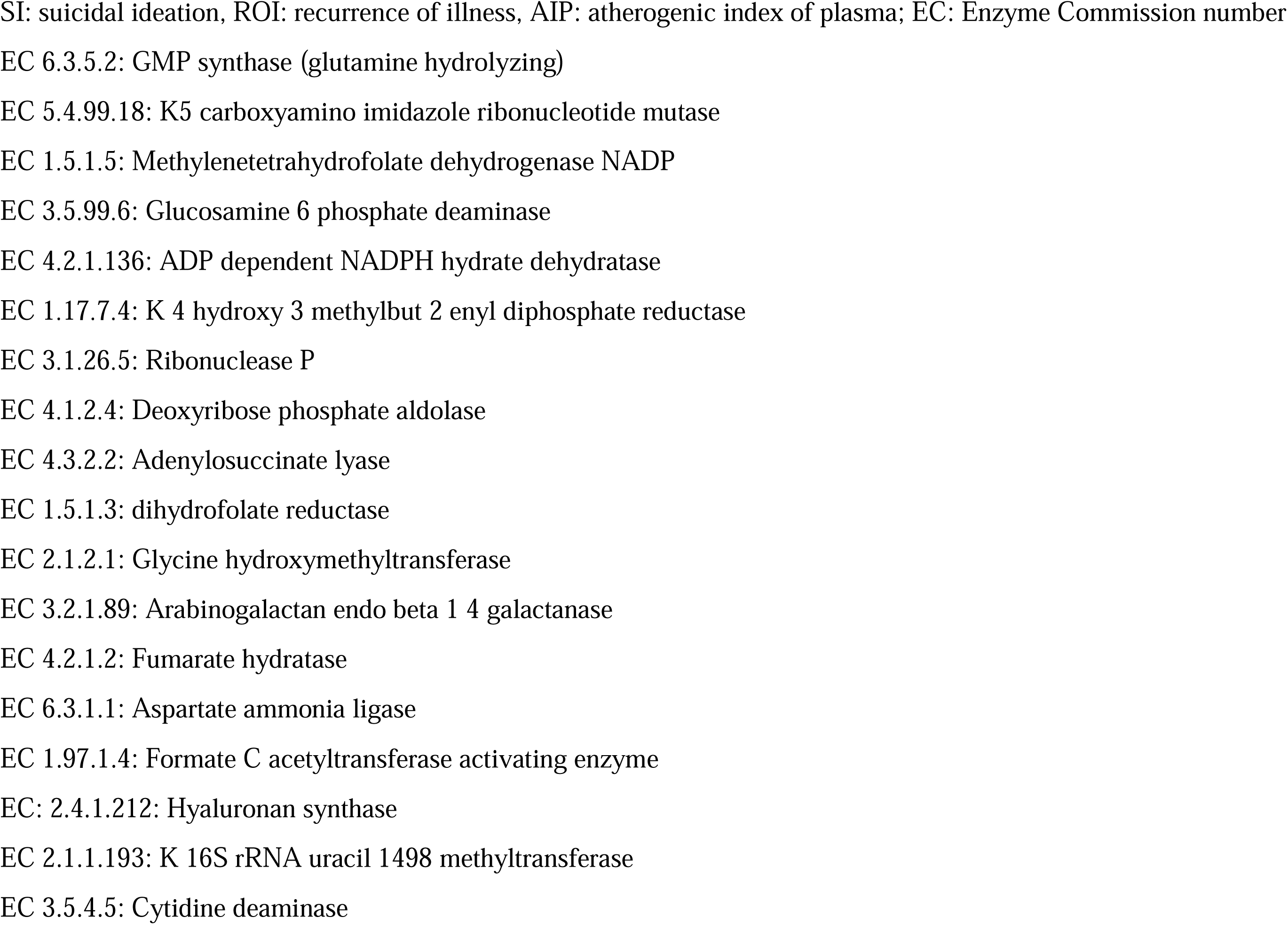
Results of multiple regression analysis with clinical data, cognitive tests scores and biomarkers as dependent variables, and enzyme families as explanatory variables.

We found that 32.2% of the variance in SI was explained by P105 – PWY TCA-cycle (positively) and PWY 7254 (inversely) (Table 2), and that 30.1% of the variance in SI (**Table 3**, regression #1) was explained by EC 6.3.5.2 and EC 4.2.1.136 (both positively). Table 2 shows that ROI is explained (15.7% of its variance) by PWY 5989. Up to 59.2% of the variance in ROI was explained by EC 1.17.7.4 and EC 4.3.2.2 (positively) and EC 3.1.26.5 and EC 4.1.2.4 (inversely). In the restricted MDD sample, ROI was positively associated with EC 1.5.1.1.3.

A large part of the variance in the Stroop tests was explained by 2 pathways (with ACEs together 59.4%) or 4 genes (56.8%). The genes and pathways were PWY 5055, EC 3.2.1.89, EC 6.3.1.1 (positively), EC 2.1.2.1, EC 4.2.1.2, and COBALSYN PWY (inversely). The growth factor composite score was predicted by 3 pathways (44.3% explained variance), namely FASYN-ELONG-PWY, LACTOSECAT PWY (positively) and PWY 6936 (inversely) and 2 genes (35.9%), namely EC 1.97.1.4 and EC 2.1.2.1(both positively). AIP levels were associated (together with BMI) with 3 genes (55.4%) and 3 pathways (57.2%). These are: EC 2.4.1.212, EC 2.1.1.193, PWY 6588 (positively), EC 3.5.4.5, PWY 6902, and P122 PWY (inversely).

We have also examined possible effects of age, sex, MetS, overweight and obesity, and the drug state of the patients on the significant taxa, genes and pathways as listed above. Fifteen patients were treated with antidepressants, three with atypical antipsychotics, and 9 with benzodiazepines. None used lithium or other mood stabilizers. We could not find any significant associations between the above demographic data and drug state and any of the taxa, genes or pathway scores that are significant in our analyses (even without *p*-correction).

## Discussion

### Gut microbiome diversity in MDD

The results of our shotgun metagenomics indicate that the overall gut microbiome diversity, regarding both species richness and evenness, is not significantly different between MDD and healthy controls. Systematic reviews demonstrate inconsistent alpha-diversity results and minimal pooled effect sizes, with considerable heterogeneity influenced by sequencing region, methodologies, geography, diet, body mass index, and psychotropic medication (Cheung et al., 2019; Gao et al., 2023; Sanada et al., 2020). Beta-diversity discrepancies identified through amplicon sequencing between MDD and controls show more prevalent alterations (Chung et al., 2019; Jiang et al., 2015; Nikolova et al., 2021; Zhang et al., 2021), though not in all studies (Maes, Vasupanrajit, Jirakran, Klomkliew, Chanchaem, Tunvirachaisakul, & Payungporn, 2023; Naseribafrouei et al., 2014). Recent extensive re-analyses validated notable β-diversity differentiation (Kim et al., 2025).

Shotgun metagenomics provides more comprehensive and reliable diversity estimates compared to short amplicon 16S, as 16S diversity can be influenced by primer selection and rRNA copy number (Durazzi et al., 2021; Madison et al., 2023; Rausch et al., 2019). Results reviewed in our Introduction endorse the utilization of shotgun metagenomics analysis where precise richness, Shannon diversity, and Bray–Curtis metrics are essential. Shotgun metagenomic analyses demonstrated moderate and somewhat uneven alterations in diversity. In cross-diagnostic research comparing MDD, bipolar depression, and controls, global α-diversity remained statistically unchanged, although compositional alterations suggested phylum-level imbalance (Rong et al., 2019). A case–control study revealed significant variations in β-diversity and pathways in MDD (Lai et al., 2021). A young-adult pilot employing rigorous confounder control indicated no global diversity disparities, although it noted species- and pathway-specific alterations (Chen et al., 2023).

### Important taxa features of MDD

Our findings show that although certain bacterial species aid in distinguishing MDD from controls, the multivariate gut microbiota profile has a small effect size. Within the species prevalent in MDD in the overall discriminant signal, *Ruminococcus gnavus, Veillonella rogosae*, and *Anaerobutyricum hallii* were recognized as substantial contributors to the MDD model, whereas *Vescimonas coprocola, Faecalibacterium prausnitzii,* and *Faecalibacillus intestinalis* abundances were significant for the control model. Our research aligns with prior findings indicating that a differentiation between MDD and control groups was achievable via either amplicon sequencing or shotgun analysis [review in (Maes, Vasupanrajit, Jirakran, Klomkliew, Chanchaem, Tunvirachaisakul, & Payungporn, 2023)]. However, the latter study indicated a lack of consistency among studies conducted in various countries and cultures. This phenomenon may be attributed not only to methodological discrepancies and technical issues, but particularly to environmental factors, such as dietary influences that significantly shape gut microbial composition (Maes, Vasupanrajit, Jirakran, Klomkliew, Chanchaem, Tunvirachaisakul, & Payungporn, 2023).

Nonetheless, there are several similarities between the present microbiota profile derived by shotgun analysis and prior reports, as all indicate a degree of compositional dysbiosis characterized by an increase in pathobionts and a reduction in probiotics. Preliminary case–control studies revealed modifications in dominant phyla and an increased occurrence of potential pathobionts in MDD (e.g., *Bacteroides, Enterobacteriaceae*), coupled with a decrease in Firmicutes in specific cohorts (Jiang et al., 2015; Naseribafrouei et al., 2014). In various studies, the most consistent finding in MDD is a relative reduction of SCFA producers, e.g., *Faecalibacterium* and *Coprococcus*, alongside an increase in taxa associated with pro-inflammatory properties or mucin utilization, aligning with hypotheses of low-grade inflammation and barrier dysfunction (Nikolova et al., 2021).

The current shotgun metagenomics investigation shows that the top 3 enriched taxa in the multivariate profile of MDD reflect a dysbiosis enterotype. *Ruminococcus gnavus* is often associated with inflammatory mechanisms that stimulate dendritic-cell TNF-α production and inflammatory bowel disease (IBD) (Hall et al., 2017; Henke et al., 2019). Furthermore, *R. gnavus* may break down mucus glycans, thereby compromising the intestinal barrier and enhancing antigen exposure (Crost et al., 2016). *Veillonella rogosae* has been associated with inflammatory markers that predict IBD relapses, suggesting context-dependent pathobiont potential (Rocha et al., 2023). *Anaerobutyricum hallii* is a Gram-positive, anaerobic bacillus that synthesizes butyrate and propionate, potentially contributing to epithelial energy, tight junction integrity, anti-inflammatory signaling, and immunomodulation (Shetty et al., 2018). Nonetheless, it may have adverse effects, including the generation of reactive aldehyde intermediates such as 3-hydroxypropionaldehyde (reuterin) and the pro-oxidant acrolein (Engels et al., 2016). Augmented generation of aldehydes and oxidative stress are significantly implicated in the NIMETOX pathophysiology of MDD (Maes et al., 2011). Furthermore, the top four (*Granulicatella*), five (*Lacrimispora amygdalina*), and six (*Blautia massiliensis*) taxa of the dysbiosis enterotype may function as pathobionts, possess limited evidence for disease association, or exacerbate pre-existing metabolic disturbances (vascular stiffness) in the host (Bokhari et al., 2025; Carlier et al., 2006; Cuadrat et al., 2023; Zilberstein et al., 2025).

Conversely, the top four protective taxa in the multivariate profile of MDD indicate a loss of the anti-inflammatory ecosystem and lower microbiome resilience. *Vescimonas coprocola*, a Gram-negative, anaerobic rod from the Ruminococcaceae family, possesses anti-inflammatory barrier-supporting properties. Secondly, *Coprococcus*, a Gram-positive, anaerobic coccus, ferments dietary fibers into SCFAs, notably butyrate (Fitzgerald et al., 2025). Third, *Faecalibacterium prausnitzii*, a Gram-negative butyrate-producing bacterium (Martín et al., 2015), exhibits a reproducible depletion (Kim et al., 2025). This bacterium augments tight junctions, fortifies barrier integrity, elevates IL-10 levels, and suppresses nuclear factor kappa-B, whereas its diminished abundance correlates with IBD (Fujimoto et al., 2013; Miquel et al., 2014; Miquel et al., 2013). Fourth, *Faecobacillus intestinalis*, a gram-positive anaerobic bacterium, exhibits probiotic potential, contributing to gut barrier integrity and SCFA synthesis (Kulecka et al., 2024).

### Pathways

A key discovery of this work is that the majority of pathways identified in MDD exhibit enrichments in pathways that potentially may aggravate NIMETOX pathways (e.g., folate transformation I, peptidoglycan biosynthesis IV, formaldehyde assimilation, and serotonin degradation (MetaCyc, 2025). The identified pathways may account for a significant portion of the variation in OSOD, suicidal ideation, and illness recurrence. For example, PWY 5989 (stearate production) is strongly associated with OSOD and ROI. This pathway favors production of saturated fatty acids and, thus, might increase lipotoxicity, which is part of the NIMETOX pathways (Maes et al., 2025).

Lai et al. previously identified modifications in microbial tryptophan biosynthesis and metabolism in MDD (Lai et al., 2021). Yang et al. integrated shotgun metagenomics with fecal metabolomics, correlating depression-related gene modules and metabolites with amino acid pathways, particularly GABA, phenylalanine/tyrosine, and tryptophan metabolism, thereby connecting microbial co-expression networks to disrupted neurotransmission and redox equilibrium (Yang et al., 2020).

The control network established in the current study reveals pathways such as catechol degradation II, NAD de novo synthesis, gluconeogenesis, and L-cysteine synthesis serving as major hubs. In addition, the control group displays considerably enhanced pathways, including the superpathway of mycolate biosynthesis, 6-hydroxymethyl-dihydropterin diphosphate biosynthesis II, and propanoate fermentation to 2-methylbutanoate, suggesting barrier-protective effects, SCFA production, and increased resistance to oxidative stress (MetaCyc, 2025).

Significantly, our research demonstrated that the neurocognitive anomalies in MDD correlate with diminished PWY 5055 scores and elevated COBALSYN PWY scores. The microbial cobalamin biosynthetic pathway may disrupt endogenous B12 levels through the creation of corrinoids, which pathogens may utilize, potentially resulting in heightened inflammation (Wexler et al., 2018). Reduced PWY 5055 (aspartate family amino acid biosynthesis) scores may lead to a deficiency of threonine, potentially resulting in compromised gut wall integrity (Faure et al., 2005).

The alterations in immune biomarkers (VEGF and SCF) in MDD are linked to three pathways. Elevated FASYN-ELONG-PWY suggests augmented microbial capacity for saturated fatty acid elongation, potentially facilitating TLR4–NF-κB signaling and NLRP3 inflammasome activation (Zhou et al., 2020). Elevated LACTOSECAT-PWY (lactose degradation I) suggests augmented microbial ability to hydrolyze and ferment luminal lactose (MetaCyc, 2025). Lactose is metabolized by this pathway by certain pathobionts, including *Staphylococcus aureus* and *Streptococcus mutans* (MetaCyc, 2025). Reduced PWY-6936 (seleno-amino acid biosynthesis) indicates the microbial ability to synthesize selenocysteine and selenomethionine analogs, as well as microbial selenoproteins (MetaCyc, 2025). Diminished antioxidant defenses, including those in selenoproteins, contribute to the NIMETOX pathophysiology of MDD (Maes et al., 2011).

We found that elevated AIP scores in MDD are largely predicted by three paths. First, elevated PWY 6588 (pyruvate fermentation to acetone) reflects heightened acetone production (MetaCyc, 2025), perhaps contributing to oxidative stress and lipid peroxidation, thus enhancing NIMETOX pathways (Bondoc et al., 1999). Second, diminished PWY 6902 (Chitin degradation II) scores signify a reduction in microbial-tolerance maintaining pathways, responsible for the synthesis of N-acetyl-D-glucosamine, which supports the gut barrier (Zhao et al., 2018), and protects against activated NIMETOX pathways (Maes, Almulla, et al., 2025). The depletion of the P122 PWY (the heterolactic fermentation pathway) suggests depletion of heterofermentative lactic acid bacteria, which possess barrier-protective and antioxidant properties, as well as the ability to cross-feed butyrate producers (Feng & Wang, 2020). A loss may thus exacerbate leaky gut, thereby aggravating the NIMETOX drift.

### Gene families

The top three primary genes associated with MDD (EC 1.1.1.86; EC 2.7.1.25; EC 4.1.1.11) indicate elevated synthesis of branched-chain amino acids (BCAAs), dephospho-CoA kinase, and β-alanine (MetaCyc, 2025). Enhancements in these microbial enzyme capacities may adversely affect the NIMETOX framework via a) exacerbating insulin resistance associated with obesity (Newgard et al., 2009) and disrupting neurotransmitter equilibrium whereby elevated BCAAs compete with L-tryptophan for blood-brain barrier transport (Maes & Meltzer, 1995); b) augmenting CoA synthesis potentially redirecting metabolism towards insulin-resistant energy pathways and mitochondrial dysfunction (Newgard et al., 2009); and c) increasing β-alanine production possibly diminishing luminal aspartate availability (Williamson & Brown, 1979), which may affect microbiome and host amino acid homeostasis. Furthermore, we discovered that 54.2% of the variance in OSOD was associated with four genes, suggesting a transition in microbial purine, amino-sugar, and one-carbon metabolism. EC 6.3.5.2 and EC 5.4.99.18 are de novo purine (GMP) biosynthesis enzymes, whereas lower EC 3.5.99.6 regulates amino-sugar metabolism (MetaCyc, 2025).

Lai et al. previously identified modifications in microbial tryptophan biosynthesis and metabolism in MDD, characterized by diminished genes for indole/serotonin production and an increase in kynurenine-related enzymes, which may promote neurotoxic kynurenines, disrupt 5-HT signaling, and activate immune responses (Lai et al., 2021). Yang et al. found correlations between depression-related gene modules and metabolites with amino acid pathways, particularly GABA, phenylalanine/tyrosine, and tryptophan metabolism, thereby connecting microbial co-expression networks to disrupted neurotransmission and redox equilibrium (Yang et al., 2020).

Conversely, gene families such as Glucosamine-6-phosphate deaminase [EC 3.5.99.6] and Deoxyribose-phosphate aldolase [EC 4.1.2.4] exhibited greater abundance in the healthy control group. Both enzymes occupy critical crossroads in microbial carbon use and their loss in stool shotgun data may indicate a decline in saccharolytic and nucleotide-scavenging processes, as well as alterations in the effective catabolism of amino-sugars (MetaCyc, 2025). Enhanced ROI, a novel pharmacological target for the treatment of recurrent MDD, was linked to elevated levels of EC 1.5.1.3 (dihydrofolate reductase), leading to the production of 5,6,7,8-tetrahydrofolate (THF) and therefore an augmented supply of one-carbon (1C) cofactors (MetaCyc, 2025). Its combination of enhanced EC 6.3.5.2 (GMP synthase, linked to suicidal ideation) may lead to an augmented production of uric acid (Ducker & Rabinowitz, 2017; Fox & Stover, 2008). Elevated blood uric acid levels have been frequently documented in MDD (Zhang et al., 2025) and exhibit a dual redox profile, which higher levels, context-dependently facilitating NIMETOX responses (Yin et al., 2019).

A substantial fraction of the variance in the immune biomarker profile (VEGF and SCF) was explained by two genes from the EC enzyme family, specifically EC 1.97.1.4 (pyruvate formate-lyase–activating enzyme) and EC 2.1.2.1 (glycine hydroxymethyltransferase). Elevated EC 1.97.1.4 suggests enhanced anaerobic formate synthesis (MetaCyc, 2025), which serves as a metabolic marker of inflammation-related dysbiosis (Hughes et al., 2017). Higher EC 2.1.2.1 produces L-serine and tetrahydromethanopterin (MetaCyc, 2025), which supports methanogenesis, favoring methane-associated constipation and slower gut transit (Escalante-Semerena et al., 1984; Ghoshal et al., 2011). Interestingly, some of the pathways linked with MDD (discussed in the previous sections) may impact gut motility, e.g., the archaetidylinositol & CDP-archaeol biosynthesis, factor 420 and PWY 2.1.2.1 (MetaCyc, 2025). These enzymes and pathways may increase methanogen abundance and methane formation, which slows intestinal transit and may worsen constipation, one key symptom in some MDD patients.

The enzyme profile linked to elevated AIP values suggests potential pro-atherogenic effects. Consequently, EC 2.4.1.212 (hyaluronan synthase) facilitates the buildup of hyaluronan in the vascular wall, potentially enhancing monocyte adhesion and inflammation (Homann et al., 2018). 16S rRNA methyltransferase genes, such as those found in *Enterobacteriaceae* and *Klebsiella*, may indicate pathobionts or their resistance mechanisms (Lioy et al., 2014). The proliferation of Enterobacteriaceae-dominated dysbiosis is associated with intestinal barrier impairment and systemic low-grade inflammation (Di Vincenzo et al., 2024).

As previously mentioned, novel shotgun-metagenomic analyses of the gastrointestinal microbiome in MDD indicate dysfunction at the microbial gene family (KEGG Ortholog) level. For instance, a case-control study examined 26 MDD patients and 29 healthy controls. The MDD group exhibited significantly lower levels of four KEGG orthologs (K01817, K11358, K01626, K01667) in the tryptophan biosynthesis/metabolism module, while K01626 exhibited an inverse correlation with HAMD depression scores (Lai et al., 2021). In a more recent study, a multi-omics adolescent cohort identified six dysregulated amino-acid-metabolism modules in fecal metagenomics, with lysine metabolism being the most evident. These modules were subsequently linked to plasma amino acids and frontal-lobe connectivity in fMRI through (Teng et al., 2025).

### Limitations

Our shotgun metagenomics analysis in patients with MDD and controls presents some limitations. The sample size of the regression analysis is somewhat limited for high-dimensional microbiome data regarding effect-size estimations. The cross-sectional approach inhibits causal inference and fails to differentiate between state and trait effects. Residual confounding from dietary and other environmental factors remains a possibility. Microbial functional profiles are derived from DNA, hence EC and PWY abundances indicate prospective metabolic activity rather than actual performance, and unclear enzyme annotations necessitated aggregation rules that may mask particular effects. Due to the compositional nature of relative-abundance data, the results are contingent upon transformation (in this case, we employed CLR and z scaling) and necessitate external replication.

## Conclusions

Our shotgun research suggests that diversity indexes exhibit no significant differences between MDD and controls. The multivariate enterotype profile observed in MDD corresponds with gut dysbiosis, a pro-inflammatory transition, barrier stress, metabolic-immune activation, and a decrease in SCFA-mediated anti-inflammatory defenses. Pathway analysis indicates loss of barrier protection, antioxidants, and SCFAs, and activation of NIMETOX pathways that may correlate with depressive symptoms, immune biomarkers, and atherogenicity in MDD individuals. MDD is accompanied by an impaired metabolic coordination, namely the microbiome in MDD is primarily concerned with regulating metabolic byproducts and LPS biosynthesis, underscoring a modified microbial milieu in MDD. The differential abundance of particular gene families indicates potential metabolic differences between MDD and controls, possibly reflecting alterations in intestinal microbial function that influence purine, sugar, and protein metabolism, gut dysbiosis and barrier stress. Therefore, it is plausible that alterations in the gut microbiome functions contribute to the peripheral NIMETOX pathophysiology of MDD. External replication in other countries and cultures is needed before the alterations in the gut microbiome could be regarded as new drug targets to treat MDD.

## Supporting information

Electronic Supplementary File

## Data availability

Data will be provided by the first author (MM) upon a reasonable request.

## Ethical statement

This study was approved by the Ethics Committee of Chulalongkorn University, Bangkok, Thailand (approval no. #62/073), and all participants gave written informed consent.

## Author’s contributions

All contributing authors have participated in the manuscript. All authors have read and agreed to the published version of the manuscript. MM designed the study. Patients were recruited by AV, KJ and CT. Microbiome assays were performed by PV, PC and SP. Statistical analyses were performed by MM, AM, and SP. AFA, YZ, SP and AM revised the paper.

