## Supplementary material for "Functional Shotgun Metagenomic Insights into Gut Microbial Pathway and Enzyme Disruptions Linking Metabolism, Affect, Cognition, and Suicidal Ideation in Major Depressive Disorder": Electronic Supplementary File

**Supplementary methods**

**Participants**

The study excluded individuals with DSM-IV-TR axis 1 diagnoses other than MDD, such as post-traumatic stress disorder, autism spectrum disorders, obsessive-compulsive disorder, bipolar disorder, schizophrenia, schizo-affective disorder, substance use disorder (excluding nicotine dependence), and psycho-organic disorders. The control group was recruited in Bangkok, Thailand, through word of mouth in the same catchment area as the patients. Controls were precluded from the study if they had a positive family history of MDD, bipolar disorder, or suicide and suffered from any of the aforementioned axis 1 disorders, dysthymia, cyclothymia, or MDD.

Moreover, individuals participating in the study, both controls and patients, were systematically excluded based on specific medical illnesses and conditions. These exclusions included: a) any allergic or inflammatory reactions occurring within three months prior to the study; b) the inclusion of pregnant or lactating women; c) the presence of medical, immune, or autoimmune disorders, which encompassed systemic lupus erythematosus, inflammatory bowel disease, cancer, chronic obstructive pulmonary disease (COPD), psoriasis, type 1 diabetes mellitus, and irritable bowel syndrome; d) individuals who had previously (lifetime) received treatment with immunomodulatory drugs, such as glucocorticoids or immunosuppressants; e) patients who had experienced moderate to critical COVID-19 or mild COVID-19 within six months prior to enrollment; f) the existence of neurological, neuroinflammatory, or neurodegenerative disorders, including stroke, Alzheimer’s disease, Parkinson’s disease, multiple sclerosis, adrenoleukodystrophy (ALD), and epilepsy; and g) subjects who had been administered antibiotics, or pharmaceutical dosages of antioxidants or omega-3 supplements within six months prior to enrollment.

Prior to engaging in this study, all participants duly submitted written informed consent. The research was carried out in compliance with both international and Thai ethical standards, as well as privacy regulations. The Institutional Review Board of the Chulalongkorn University Faculty of Medicine, located in Bangkok, Thailand, designated as #528/63, has granted approval for the study. This approval aligns with the International Guidelines for the Protection of Human Subjects, as stipulated by the Declaration of Helsinki, The Belmont Report, the CIOMS Guideline, and the standards set forth by the International Conference on Harmonization in Good Clinical Practice (ICH-GCP).

**Clinical assessments**

A research psychologist, well-versed in the examination of affective disorders, undertook semi-structured interviews to gather socio-demographic and clinical data. This included an assessment of medical history, the frequency of prior depressive episodes, familial medical background, and the utilization of psychotropic medications. A senior psychiatrist diagnosed MDD by employing the criteria outlined in the DSM-5 and utilizing the Mini International Neuropsychiatric Interview (M.I.N.I.) as referenced by Udomratn and Kittirattanapaiboon et al. (2004). The M.I.N.I. was employed to assess axis-1 diagnoses and to appropriately exclude both patients and controls from the study. The M.I.N.I. was employed to assess Axis I diagnoses and to appropriately exclude both patients and controls from the study. The 17-item Hamilton Depression Rating Scale (HAMD) was employed by the research psychologist to evaluate the intensity of depressive symptoms(Hamilton, 1960). The Beck Depression Inventory II was employed to evaluate the intensity of self-reported depressive symptoms(Beck et al., 1996). The latter constitutes a 21-item self-report inventory, which was translated into Thai by Nantika Thavichachart and colleagues for the purpose of evaluating both the presence and severity of depressive symptoms(Thavichachart et al., 2009). The state version of the State-Trait Anxiety Inventory (STAI) was employed to evaluate state anxiety, as outlined by Spielberger et al. (1983). The Columbia Suicide Severity Rating Scale (C-SSRS) was employed to evaluate the severity of both lifetime and current suicidal ideation and attempts. The C–SSRS was developed by Posner and colleagues(Posner et al., 2011). The assessment evaluates the degree and intensity of suicidal thoughts, attempts, lethality, and self-injurious behavior that occurs without suicidal intent. We conducted a calculation of the first principal components derived from lifetime suicidal ideation (LT_SI), LT suicide attempts (LT_SA), and current SI items, as previously elucidated(Maes et al., 2024). Consequently, we extracted a principal component from the HAMD, BDI, STAI, and current_SI data, labeled overall severity of depression (OSOD)(Maes et al., 2024). The ROI index was derived by extracting the first principal component from the total number of depressive episodes, LT_SI, and LT_SA(Maes et al., 2024).

The research psychologist conducted an examination of the Stroop color and word test, specifically focusing on its three components: part 1, which serves as a neutral trial to assess reaction times; part 2, which involves a congruent trial; and part 3, which entails an incongruent trial (Stroop, 1935). We computed a z composite score based on those three test results indicating an integrated index of Stroop test results. The measurement of Adverse Childhood Experiences (ACEs) was conducted utilizing a Thai translation of the Adverse Childhood Experiences Questionnaire(Rungmueanporn et al., 2019). In the current investigation, we employed five domains of the ACE framework, which encompass sexual abuse, emotional abuse, physical abuse, emotional neglect, and physical neglect. We computed a z composite score based on the 4 last ACE items (labeled as ACEs) and used this together with the sexual abuse score in the analyses. The diagnosis of Metabolic Syndrome (MetS) was conducted in accordance with the criteria set forth by the International Diabetes Federation(Alberti et al., 2006). The calculation of body mass index (BMI) involves dividing an individual’s weight, measured in kilograms, by the square of their height, expressed in meters. The identification and diagnosis of tobacco use disorder (TUD) were conducted in accordance with the criteria established by the DSM-5.

**Blood biomarkers**

In the early morning hours, particularly between 8:00 and 9:00 a.m., a volume of 5 ml of venous blood was procured from each participant following a fasting period. The procedure for blood collection was executed utilizing a disposable syringe. The blood samples were preserved at -80°C until they were subsequently thawed for biomarker testing. As previously elucidated(Maes et al., 2024), we assessed the serum levels of VEGF and SCF utilizing Bio-Plex Multiplex Immunoassays (Bio-Rad Laboratories Inc., USA). As explained previously, we computed the ratio between both growth factors as a composite z VEGF – z SCF(Maes et al., 2024). The concentrations of high-density lipoprotein cholesterol (HDLc) and triglycerides were assessed utilizing the Alinity C system, developed by Abbott Laboratories (Otawara-Shi, Tochigi-Ken, Japan). This analysis employed an accelerator selective detergent for the measurement of HDL cholesterol and glycerol phosphate oxidase for triglyceride quantification. The coefficients of variation for HDL and triglycerides were observed to be 2.6% and 2.2%, respectively. In our investigation, we employed a composite score based on z units that represents the atherogenic index of plasma (AIP), calculated as z triglycerides minus z HDLc(Morelli et al., 2021; Mousa et al., 2022).

**Assays**

**DNA Extraction and shotgun metagenomic sequencing**

DNA was extracted from fecal samples (approximately 20 mg) using ZymoBIOMICS™ DNA Miniprep Kit (ZYMO Research, USA) following the manufacturer’s protocol. Then, 100 ng of extracted DNA were used for DNA library construction using MGIEasy FS DNA Library Prep Set (MGI, China) according to the manufacturer’s recommended protocol. After that, DNA libraries were sequenced with paired-end (2 x 150 bp) using the DNBSEQ-G400 platform (MGI, China) following the manufacturer’s recommendations.

**Metagenomic Sequence Data Processing and Differential Abundance Analyses**

Raw paired-end sequencing reads underwent quality control to ensure reliable microbial data. FastQC v0.11.9 (https://github.com/s-andrews/FastQC) was used to evaluate sequence metrics such as GC content, base quality, and adapter contamination, followed by Fastp v0.20.1 (https://github.com/OpenGene/fastp) to trim low-quality bases (Phred score <30), remove adapters, and discard reads shorter than 60 base pairs. To remove human DNA contamination, reads were aligned to the hg38 genome using BWA v0.7.18 (https://github.com/lh3/bwa), and Samtools v1.20 (https://github.com/samtools/samtools) was used to filter out human reads, leaving microbial reads for downstream analysis.

Taxonomic profiling was performed using MetaPhlAn v4.0.6 (https://github.com/biobakery/MetaPhlAn), which relies on clade-specific marker genes from the mpa_vJun23_CHOCOPhlAnSGB_202307 database. Only species with a relative abundance greater than 0.1% in at least five samples were retained. Functional profiling was conducted using HUMAnN v3.9 (https://github.com/biobakery/humann), aligning microbial reads to species-specific pangenomes via Bowtie2 v2.5.4 (https://github.com/BenLangmead/bowtie2), followed by alignment to the UniRef90 v201901 protein database using DIAMOND v2.1.9 (https://github.com/bbuchfink/diamond).

**Statistics**

Functional abundances, including gene families and pathways, were normalized to reads per kilobase (RPK) to account for sequencing depth variability. The data pertaining to microbiota, genes and pathways underwent a centered log-ratio (CLR) transformation, yielding CLR abundance values and gene and pathway scores. Taxonomic abundance analysis was conducted using the vegan R package v2.5-7 (https://github.com/vegandevs/vegan). Alpha diversity metrics, such as Shannon diversity and observed richness, were compared between MDD and HC groups using the Mann-Whitney U test, while beta diversity was assessed through Bray-Curtis dissimilarity and visualized with PCoA plots. Hierarchical clustering using hclust2 (https://github.com/SegataLab/hclust2) was employed to identify distinct clustering patterns between MDD and HC samples. A Random Forest model was implemented using the caret and randomForest packages (https://github.com/topepo/caret, https://github.com/cran/randomForest) to classify groups based on species abundance, with taxa importance ranked by the Mean Decrease in Gini index. For gene family abundance, a Ridge regression model was applied using SIAMCAT (https://github.com/zellerlab/siamcat) to identify gene families distinguishing MDD from HC groups. Model validation was performed using ROC and Precision-Recall curves, while heatmaps visualized the top gene families contributing to group separation. Pathway abundance was analyzed by calculating log2 fold changes between MDD and HC groups, with results visualized via ggplot2 (https://github.com/tidyverse/ggplot2). Spearman correlation networks were generated using igraph (https://github.com/igraph/igraph), visualized using ggraph (https://github.com/thomasp85/ggraph), and central pathways were identified based on Eigenvector Centrality analysis.

The analysis of contingency tables was utilized to examine associations among variables based on categorical data, employing Chi-square testing as a methodological approach. An analysis of variance was employed to evaluate the differences among the distinct study groups concerning continuous variables, which encompass biomarkers and clinical data. In conjunction with the manual multiple regression approach, we implemented automated regression techniques to determine the gene and pathway scores that serve as predictors for the phenome and biomarker characteristics. In this investigation, we performed a ridge regression analysis employing a regularization parameter of λ=0.1 and establishing a tolerance level at 0.4, utilizing Statistica, Windows version 12. Furthermore, we utilized forward stepwise automatic linear modeling analyses, applying an overfit criterion for both the inclusion and exclusion of variables, with a maximum effects threshold set at 5 (utilizing SPSS, Windows version 30). We conducted the best subsets analysis employing a criterion for overfit prevention, concentrating on the 10 most significant microbiota, genes or pathways identified in the previously mentioned regressions, which were executed utilizing SPSS version 30. Following these analyses, we performed a manual regression analysis using SPSS 30 and conducted a comprehensive analysis of the model statistics, which included the F statistic, degrees of freedom, and p-values, alongside the total variance explained by R². Furthermore, we undertook a comprehensive analysis of the standardized beta coefficients associated with each predictor, in conjunction with their respective t statistics and precise p-values. The final models were checked for multivariate normality (including the residual distributions and P-P plots). An evaluation of the variance inflation factor and tolerance was performed to determine any potential concerns associated with collinearity or multicollinearity. The assessment of heteroskedasticity was conducted utilizing both the White test and the modified Breusch-Pagan test to ascertain homoscedasticity. Furthermore, we conducted and elucidated a partial regression analysis of clinical data in association with the gene and pathway scores. The statistical analyses that have been referenced were conducted utilizing IBM SPSS, Windows version 30. The significance level for all statistical analyses was set at 0.05, utilizing two-tailed tests.

The sample size calculation was based on the findings reported by Maes et al(Maes et al., 2024), who reported that a large part of the variance in OSOD (namely 45.8%), which serves as the primary outcome measure, could be predicted by the multivariable regression on 5 different taxa. Using G*Power 3.1.9.4 software, power calculation was completed for the primary statistical analysis in this study, namely multiple regression analysis with an effect size of 0.53 (that is around 35% in the phenome explained), an alpha level of 0.05 (two-tailed), a power of 0.8, and five covariates maximal. The power analysis showed a minimum required sample size for this analysis of 30 given the established effect size.
